# Epigenetic entropy, social disparity, and health and lifespan in the Women’s Health Initiative

**DOI:** 10.1101/2025.02.21.25322696

**Authors:** Khyobeni Mozhui, Athena Starlard-Davenport, Yangbo Sun, Aladdin H. Shadyab, Ramon Casanova, Fridtjof Thomas, Robert B Wallace, Jay H Fowke, Karen C Johnson

## Abstract

The pace of aging varies between individuals and is marked by changes in DNA methylation (DNAm) including an increase in randomness or entropy. Here, we computed epigenetic scores of aging and entropy using DNAm datasets from the Women’s Health Initiative (WHI). We investigated how different epigenetic aging metrics relate to demographic and health variables, and mortality risk. Income and education, two proxies of socioeconomics (SE), had consistent associations with epigenetic aging and entropy. Notably, stochastic increases in DNAm at sites targeted by the polycomb proteins were significantly related to both aging and SE. While higher income was associated with reduced age-related DNAm changes in White women, the protective effect of income was diminished in Black and Hispanic women, and on average, Black and Hispanic women had relatively more aged epigenomes. Faster pace of aging, as estimated by the DunedinPACE, predicted higher mortality risk, while the maintenance of methylation at enhancer regions was associated with improved survival. Our findings demonstrate close ties between social and economic factors and aspects of epigenetic aging, suggesting potential biological mechanisms through which societal disparities may contribute to differences in health outcomes and lifespan across demographic groups.

## INTRODUCTION

Epigenetic clocks such as the Horvath, Hannum, PhenoAge, and DunedinPACE are machine-learning (ML) based predictive models that estimate the biological age, or the rate-of-aging, of an individual.^1–4^ The underlying biomolecular data is DNA methylation (DNAm), the epigenetic process that entails a chemical modification to the DNA—specifically, whether a cytosine residue that is adjacent to a guanine (a CpG site) has a methyl- tag or is unmethylated. These epigenetic markers can be measured from readily accessible biofluids such as blood or saliva and are informative of whether a person is aging faster or slower relative to their chronological age. The first-generation epigenetic clocks (e.g., Horvath, Hannum) are accurate predictors of chronological age because the algorithms were mainly trained on the age variable.^1,2^ The second-generation clocks (e.g., PhenoAge, GrimAge) were trained on a broader panel of age-related health parameters and perform remarkably well at assessing overall health, disease states, and life expectancy.^3,5,6^ The more recent DunedinPACE model was developed by tracking longitudinal changes in health traits from age-matched individuals over decades, and it directly measures the pace-of-aging.^4^

These existing models estimate biological age using preselected sets of CpGs that were picked by the respective ML models and assigned specific weights, and the final value of aging is a weighted combination of methylation levels at these distinct CpGs sites. The lists of preselected CpGs are typically unique for each model, and it is unclear as to why the “mind of the algorithm” selected those specific sites and were assigned those weights.^7–9^ While there are now several such ML-based biomarkers that are widely utilized in epidemiological research, the mechanistic pathways and the extent to which these represent stochastic versus programmatic processes of aging remain largely undefined.^10,11^ However, several of the DNAm clocks do share some common themes that hint at the underlying biology. For instance, relative to the genome-wide background, bivalent chromatin states and DNA sequences bound by the polycomb repressive complex (particularly PRC2) are consistently overrepresented among both the first- and second-generation clocks.^3,6,12^ The bivalent and PRC2 bound sequences are evolutionarily conserved and play critical roles during embryonic development and cell fate determination.^13^ A common feature of aging is for CpGs in these chromatin states to drift from a low methylation to a high methylation state, and the gain in methylation with aging has been implicated in cancers and other health risks.^14^ This is closely related to the concept of epigenetic entropy, a term borrowed from Shannon’s Information theory, that refers to the level of randomness that can be computed from DNAm data either at the global genome-wide scale, or at regional scale.^14–16^ These stereotypical shifts in the epigenetic landscape and the accumulation of stochastic variability likely contribute to some of the signal captured by the DNAm aging biomarkers.^9,14,16,17^

The Women’s Health Initiative (WHI) is a long-term prospective study of postmenopausal women that began recruitment in 1993 and has been collecting extensive health, medical, and lifespan data since that time.^18–20^ Previous epigenetic studies of DNAm clocks in the WHI have shown that a higher rate of epigenetic aging is predictive of lifespan. Prior studies also report associations with lung cancer,^21^ insomnia and immune aging,^22^ cognitive impairment and dementia,^23^ diabetes related traits, and cardiovascular health.^24–27^ Notably, the second-generation PhenoAge clock has strong associations with social disparities, and WHI participants with lower education exhibited more advance epigenetic aging. Liu et al. also found that an accelerated PhenoAge partly explain the disparity in life expectancy between racial and ethnic groups.^27^ There is growing evidence that these epigenetic biomarkers not only tell us about the intrinsic aging of cells but are strongly influenced by the larger social and environmental context. In the United States, recent studies in young as well as older adults have shown that the DunedinPACE detects more rapid aging among Black participants and among participants with low education and income levels.^28,29^ This sensitivity to social factors should not be entirely surprising since variables such as education, income inequity, and experiences of societal biases have profound and long-lasting impact on health, stress, and mental and emotional well-being, and are likely linked to the differential rates of biological aging. In fact, these close ties between the epigenetic models of aging and social variables have been interpreted in light of the “weathering hypothesis”, which proposes that the chronic exposure to socioeconomic disadvantage, and to race-based and other stressors lead to accelerated aging and higher disease burden among African Americans and other marginalized groups.^27–30^

However, a caveat to keep in mind is that these ML-based epigenetic biomarkers were initially trained in cohorts that were predominantly, or in the case of the DunedinPACE, almost exclusively of European ancestry.^4^ The training conditions likely introduce some algorithmic biases and may not generalize in full to other populations. In the present work, we attempt to unlink the epigenetic readouts from potential training-based effects by computing measures of epigenetic entropy and variability in CpG methylation that do not rely on training algorithms. For the present study, the only pre-selection or sub-setting of CpGs we performed was based on biologically informed chromatin states. The questions we ask are: (1) *how do these readouts of epigenetic entropy and stochasticity, and gross chromatin states relate to the training-based models of aging*? And, *are these non-ML readouts as informative of health and socioeconomic variables as the ML-based models, and predictive of life expectancy*? Notably, we present novel evidence linking measures of epigenetic stochasticity with social and racial disparities. Furthermore, we uncover intriguing connections between methylation maintenance at active enhancer regions and lifespan.

## RESULTS

### Datasets and participant characteristics

The present work is a secondary analysis of DNAm data generated by three WHI ancillary studies: (1) WHI-EMPC/AS315, ^26,31^ (2) BA23, ^21,32^ and (3) AS311.^33^ All three datasets measured blood DNA methylation using the Illumina Infinium Methylation 450K BeadChip (Illumina Inc.). Brief summaries of baseline characteristics are provided in **Table 1**, and sample inclusion/exclusion chart is in **Fig S1**. BA23 and AS311 used blood collected at the time of eligibility screening visit (SV) and prior to randomization to a study arm in the clinical trials (CT) or enrollment in the observational study (OS). In AS315, DNA methylation was measured either at SV (64%), annual visit 3 (29%), or annual visit 6 (7%) in the CT only. We used the larger and more diverse AS315 dataset to describe the large-scale topology of the methylome, and all analyses were initially performed in AS315 (including a sensitivity analysis using only blood collected at the SV). The main findings were then tested for replication in BA23 and AS311 after excluding participants who were also part of AS315 (**Fig S1**).

**Table 1.**
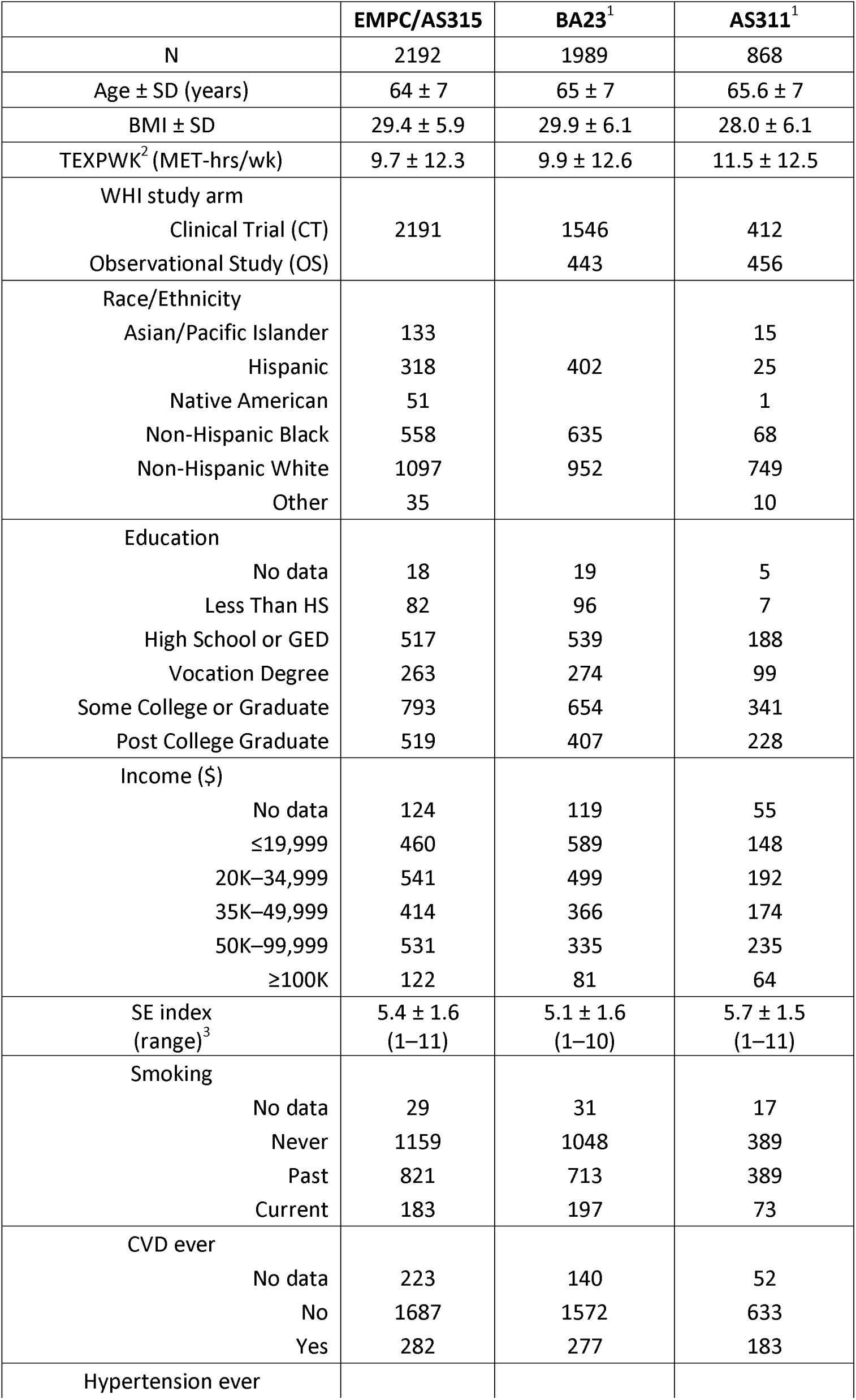
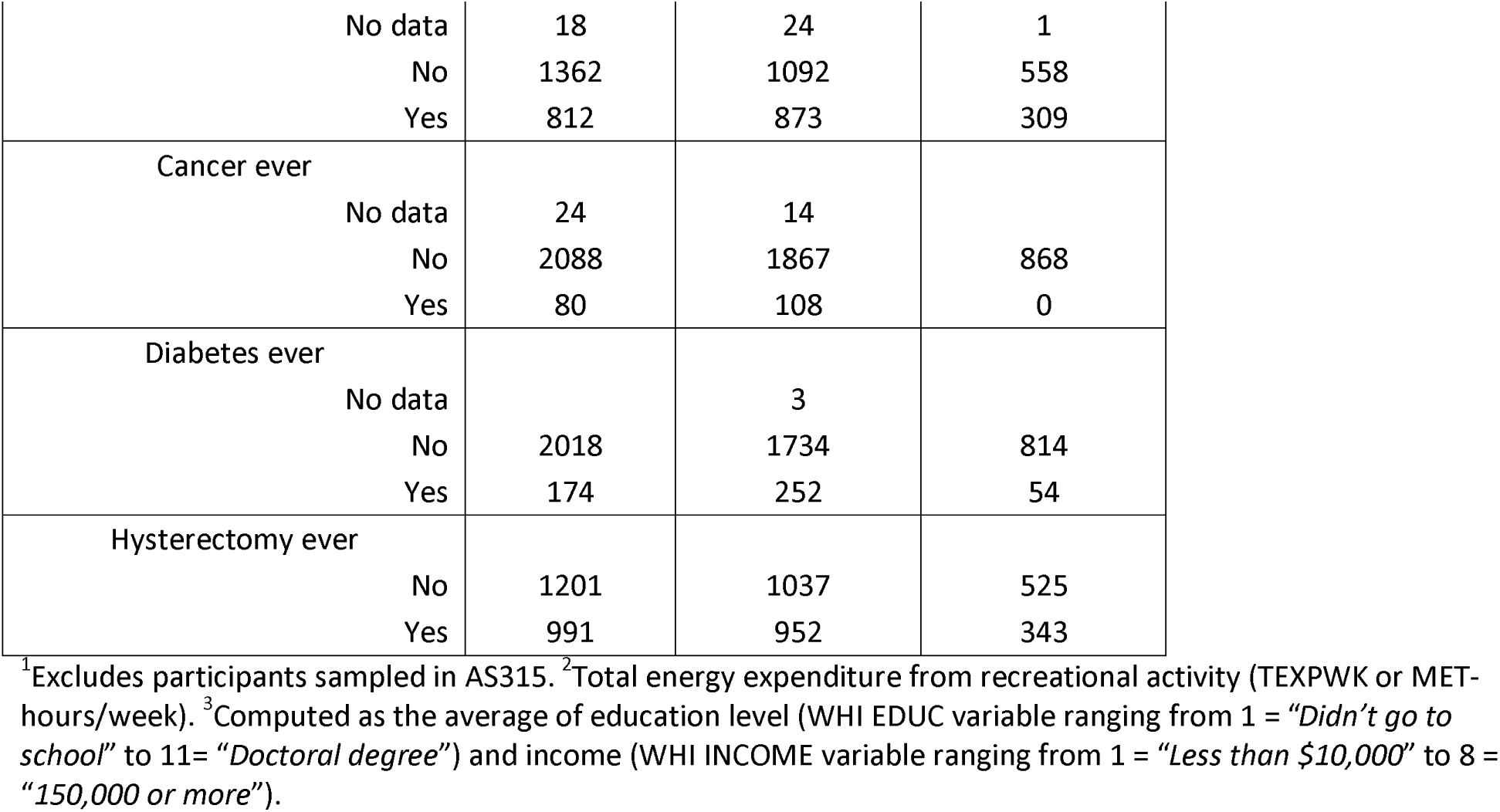
Datasets and baseline characteristics.

|  | EMPC/AS315 | BA23<sup>1</sup> | AS311<sup>1</sup> |
| --- | --- | --- | --- |
| N | 2192 | 1989 | 868 |
| Age ± SD (years) | 64 ± 7 | 65 ± 7 | 65.6 ± 7 |
| BMI ± SD | 29.4 ± 5.9 | 29.9 ± 6.1 | 28.0 ± 6.1 |
| TEXPWK<sup>2</sup> (MET-hrs/wk) | 9.7 ± 12.3 | 9.9 ± 12.6 | 11.5 ± 12.5 |
| WHI study arm |  |  |  |
| Clinical Trial (CT) | 2191 | 1546 | 412 |
| Observational Study (OS) |  | 443 | 456 |
| Race/Ethnicity |  |  |  |
| Asian/Pacific Islander | 133 |  | 15 |
| Hispanic | 318 | 402 | 25 |
| Native American | 51 |  | 1 |
| Non-Hispanic Black | 558 | 635 | 68 |
| Non-Hispanic White | 1097 | 952 | 749 |
| Other | 35 |  | 10 |
| Education |  |  |  |
| No data | 18 | 19 | 5 |
| Less Than HS | 82 | 96 | 7 |
| High School or GED | 517 | 539 | 188 |
| Vocation Degree | 263 | 274 | 99 |
| Some College or Graduate | 793 | 654 | 341 |
| Post College Graduate | 519 | 407 | 228 |
| Income (\$) | | | |
| No data | 124 | 119 | 55 |
| ≤19,999 | 460 | 589 | 148 |
| 20K–34,999 | 541 | 499 | 192 |
| 35K–49,999 | 414 | 366 | 174 |
| 50K–99,999 | 531 | 335 | 235 |
| ≥100K | 122 | 81 | 64 |
| SE index (range)<sup>3</sup> | 5.4 ± 1.6 (1–11) | 5.1 ± 1.6 (1–10) | 5.7 ± 1.5 (1–11) |
| Smoking |  |  |  |
| No data | 29 | 31 | 17 |
| Never | 1159 | 1048 | 389 |
| Past | 821 | 713 | 389 |
| Current | 183 | 197 | 73 |
| CVD ever |  |  |  |
| No data | 223 | 140 | 52 |
| No | 1687 | 1572 | 633 |
| Yes | 282 | 277 | 183 |
| Hypertension ever |  |  |  |

|  |  |  |  |  |
| --- | --- | --- | --- | --- |
|  | No data | 18 | 24 | 1 |
|  | No | 1362 | 1092 | 558 |
|  | Yes | 812 | 873 | 309 |
| Cancer ever |  |  |  |  |
|  | No data | 24 | 14 |  |
|  | No | 2088 | 1867 | 868 |
|  | Yes | 80 | 108 | 0 |
| Diabetes ever |  |  |  |  |
|  | No data |  | 3 |  |
|  | No | 2018 | 1734 | 814 |
|  | Yes | 174 | 252 | 54 |
| Hysterectomy ever |  |  |  |  |
|  | No | 1201 | 1037 | 525 |
|  | Yes | 991 | 952 | 343 |

<sup>1</sup>Excludes participants sampled in AS315. <sup>2</sup>Total energy expenditure from recreational activity (TEXPWK or MET- hours/week). <sup>3</sup>Computed as the average of education level (WHI EDUC variable ranging from 1 = "Didn't go to school" to 11= "Doctoral degree") and income (WHI INCOME variable ranging from 1 = "Less than \$10,000" to 8 = "150,000 or more").

### Epigenetic stochasticity and how it relates to chromatin states and aging

The methylation beta-values show the expected bimodal distribution with most CpGs at beta-values close to 0 (i.e., most cells are unmethylated at that CpG) or 1 (most cell are methylated at that CpG) (**Fig 1a**). With aging, cells begin to drift from their initial methylation states,^34^ and this results in an increase in entropy and a shift towards a beta = 0.5 (a “hemi-methylated” state). We computed the methylome-wide entropy using all CpGs that had complete data in all participants. To illustrate the “landscape erosion” towards a hemi-methylated and presumably more random state (i.e., beta ∼ 0.5), **Fig 1a** displays 5 participants with Shannon entropy > 0.85, and 5 with entropy <0.4. This global measure of epigenetic discordance showed a wide variation in the AS315 cohort (**Fig 1b**) but had only a weak positive correlation with age (r=0.08, p = 0.0002; **Fig 1c**).

**Fig 1.**
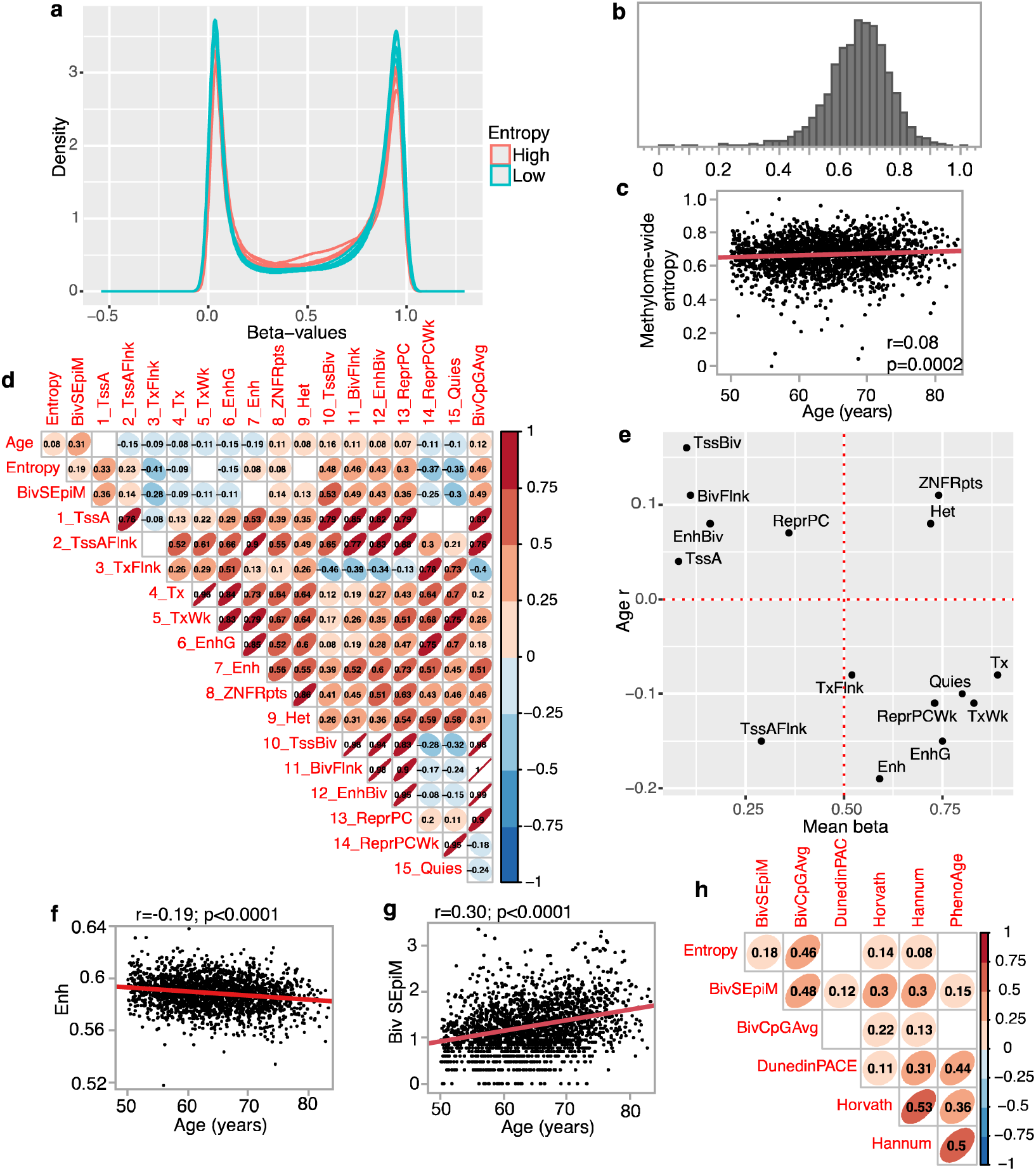
(a) Density plots of methylome-wide beta-values for 10 AS315 participants (all identified as non-Hispanic White). 5 have entropy ≤0.4 (cyan), and 5 have entropy ≥0.85 (soft red). (b) Histogram of methylome-wide entropy shows wide variability in AS315. (c) Weak but positive correlation between chronological age and methylome entropy in AS315. (d) Pair-wise correlations between age and the different non-training based epigenetic readouts including mean methylation by chromatin states (due to the large number of comparisons, the graph only displays correlations with p < 0.001). (e) The x-axis is the mean beta-values for the 15 chromatin states; y-axis is the Pearson r between these states and chronological age. Mean methylation at enhance CpGs decrease (f), while levels of stochastic epimutations at the three bivalent sites (TssBiv, BivFlnk, EnhBiv) increase with chronological age (g). (h) Pair-wise correlation for the non-training based measures (overall entropy, stochasticity at bivalent sites, and average beta-values for the bivalent CpGs) and four train-based measures of aging (only displays correlations with p < 0.001). All these are residual values adjusted for chronological age.

As methylation levels are highly dependent on the genomic context, we annotated each CpG for the predicted chromatin state based on the Roadmap Epigenomics Consortium’s ChromHMM 15-states model.^35–38^ This overlays the DNAm data with histone-based predicted epigenetic states and regulatory factors (**Table 2**).^35^ For each participant, we computed the average beta-values at each of the 15 states thereby reducing the ∼450,000 features to 15 values. The mean methylation at the chromatin states showed varying levels of correlation with chronological age (**Fig 1d**; **Table 2**). The highest positive correlate of age was the repressed bivalent transcription start site (TssBiv; r = 0.16, **Fig S2a**), and the highest negative correlate was for enhancer regions (Enh; r = -0.19, **Fig 1f**).

**Table 2.** Predicted chromatin states for the >450,000 CpG.

| ChromHMM | Counts<sup>1</sup> | Repressed/<br>Active | Description | Non-coding<br>conserved elements | Mean beta<br>in AS315 | r<br>(age)<sup>2</sup> | p<br>(age)<sup>2</sup> |
| --- | --- | --- | --- | --- | --- | --- | --- |
| 1_TssA | 124139 | Active | Active TSS | Strongly enriched | 0.08 | 0.04 | 0.0476 |
| 2_TssAFlnk | 23774 | Active | Flanking Active TSS | Strongly enriched | 0.29 | -0.15 | <.0001 |
| 3_TxFlnk | 1001 | Active | Transcribed state at gene 5' and 3' | Moderately<br>enriched | 0.52 | -0.08 | <.0001 |
| 4_Tx | 32162 | Active | Strong transcription | No enrichment | 0.89 | -0.08 | 0.0001 |
| 5_TxWk | 50018 | Active | Weak transcription | No enrichment | 0.83 | -0.11 | <.0001 |
| 6_EnhG | 1545 | Active | Genic enhancers | Moderately<br>enriched | 0.75 | -0.15 | <.0001 |
| 7_Enh | 20980 | Active | Enhancers | Strongly enriched | 0.59 | -0.19 | <.0001 |
| 8_ZNF/Rpts | 987 | Active | ZNF genes & repeats | No enrichment | 0.74 | 0.11 | <.0001 |
| 9_Het | 3468 | Repressed | Heterochromatin | No enrichment | 0.72 | 0.08 | 0.0002 |
| 10_TssBiv | 6833 | Repressed | Bivalent/Poised TSS | Strongly enriched | 0.10 | 0.16 | <.0001 |
| 11_BivFlnk | 8290 | Repressed | Flanking Bivalent TSS/Enh | Strongly enriched | 0.11 | 0.11 | <.0001 |
| 12_EnhBiv | 4869 | Repressed | Bivalent Enhancer | Strongly enriched | 0.16 | 0.09 | <.0001 |
| 13_ReprPC | 32422 | Repressed | Repressed PolyComb | Strongly enriched | 0.36 | 0.07 | 0.0007 |
| 14_ReprPCWk | 33551 | Repressed | Weak Repressed PolyComb | Weakly enriched | 0.73 | -0.11 | <.0001 |
| 15_Quies | 136857 | Repressed | Quiescent/Low | No enrichment | 0.80 | -0.10 | <.0001 |
| NA | 1339 |  |  |  |  |  |  |

<sup>1</sup>Number of CpG probes on the Illumina 450K platform located in these predicted chromatin states. <sup>2</sup>Pearson correlation and p with chronological age in AS315

Whether a CpG positively or negatively correlates with chronological age is partly dependent on the average methylation of its chromatin state, and this is depicted in **Fig 1e**. For instance, CpGs at the bivalent sites—TssBiv, BivFlnk, and EnhBiv— are known to have low mean methylation and gain methylation with aging, and this pattern is also seen in the WHI (plots to the top left quadrant of **Fig 1e**).^39^ These bivalents states also had the highest positive r with entropy (e.g., **Fig S2c**). For this reason, we explored the idea of “stochastic epimutations” (SEpiM) at these sites. We implemented an outlier detection approach,^40,41^ and counted for positive outliers at TssBiv, BivFlnk, and EnhBiv. As these CpGs gain methylation with aging, a positive outlier with higher beta-value is presumed to represent a more aged methylome. The outlier counts (what we refer to as BivSEpiM) showed strong positive correlation with chronological age (**Fig 1g**). In contrast to the bivalent sites, CpGs that flank actively transcribed genes (TxFlnk) had mean methylation of beta-value ∼0.5 and had the strongest negative correlation with entropy **(Fig S2d**) and had modest inverse correlation with age (bottom right quadrant in **Fig 1e**). Weakly repressed polycomb (ReprPCWk) and Quiescent (Quies) were also regions with higher mean methylation (beta-values >0.70) and negatively correlated with both entropy and age. Collectively, the results indicate that CpGs in low methylation sites (e.g., TssA, TssBiv, etc.) gain methylation with aging, and contribute to higher entropy. In contrast, CpGs in sites with higher mean methylation (TxFlnk, ReprPCWk and Quies) tend to lose methylation with aging, and methylation maintenance at these sites may contribute to lower entropy.

### Relating epigenetic stochasticity to training-based DNAm biomarkers

To evaluate how the non-training based DNAm readouts relate to the training-based models of biological aging, we compared pace or rates of biological aging (i.e., age-deviation or age-acceleration) estimated by the original Horvath and Hannum clocks, and the newer PhenoAge and DunedinPACE.^1–4^ After adjusting for chronological age, higher BivSEpiM was consistently associated with a higher pace of aging as defined by the prior models (**Fig 1h**). The methylome-wide entropy was positively correlated with higher age-acceleration as estimated by the Horvath or Hannum clocks (**Fig 1h**). This indicates that the overall buildup of discordance in the methylome partly, but not fully, contributes to these training-based biomarkers of aging.

### Epigenetic stochasticity and social and health variables at baseline

We treated the age-regressed residuals of the DNAm readouts (i.e., age-deviation) as outcome variables, and explored associations with baseline health and social variables. For rigor, we performed the analysis with multiple regression models in the AS315 that included a sensitivity analysis; for nominally significant associations (uncorrected p ≤ 0.05), we tested for replication in BA23 and AS311. Regarding the interpretation of age-deviation, as all the DNAm readouts listed in **Table 3** increase in value with aging, a positive age-deviation indicates higher rate of aging, and negative indicates slower rate of aging. Model 1 was a multivariable regression with race/ethnicity, socioeconomic index (SE index, a combination of self-reported income and education), smoking status, body weight index (BMI), and total weekly recreation energy expenditure (TEXPWK; MET-hours/week) as predictors. To make the magnitudes of effects comparable, **Fig 2a** displays the standardized regression estimates. For the three non-training-based readouts (i.e., entropy, BivSEpiM, and BivCpGAvg), the most consistent association was with SE index (**Table 3**). SE index also had significant negative associations with the DunedinPACE and the PhenoAge clock (**Fig 2a**; unstandardized regression estimates in **Data S1**.). A unit increase in SE index resulted in -0.004 lower entropy, -0.02 lower BivSEpiM, and -0.005 slower DunedinPACE of aging (**Table 3**). The epigenetic clocks, which are in units of years provide a more intuitive interpretation. For the PhenoAge, one unit increase in SE index resulted in roughly 0.25 years lower biological age relative to chronological age. We note that the bivalent readouts and DunedinPACE showed higher aging rates in all other race/ethnicity groups relative to White participants. Consistent with previous reports, the Hannum clock detected a significantly slower rate of aging among the Black participants.^42^

**Fig 2.**
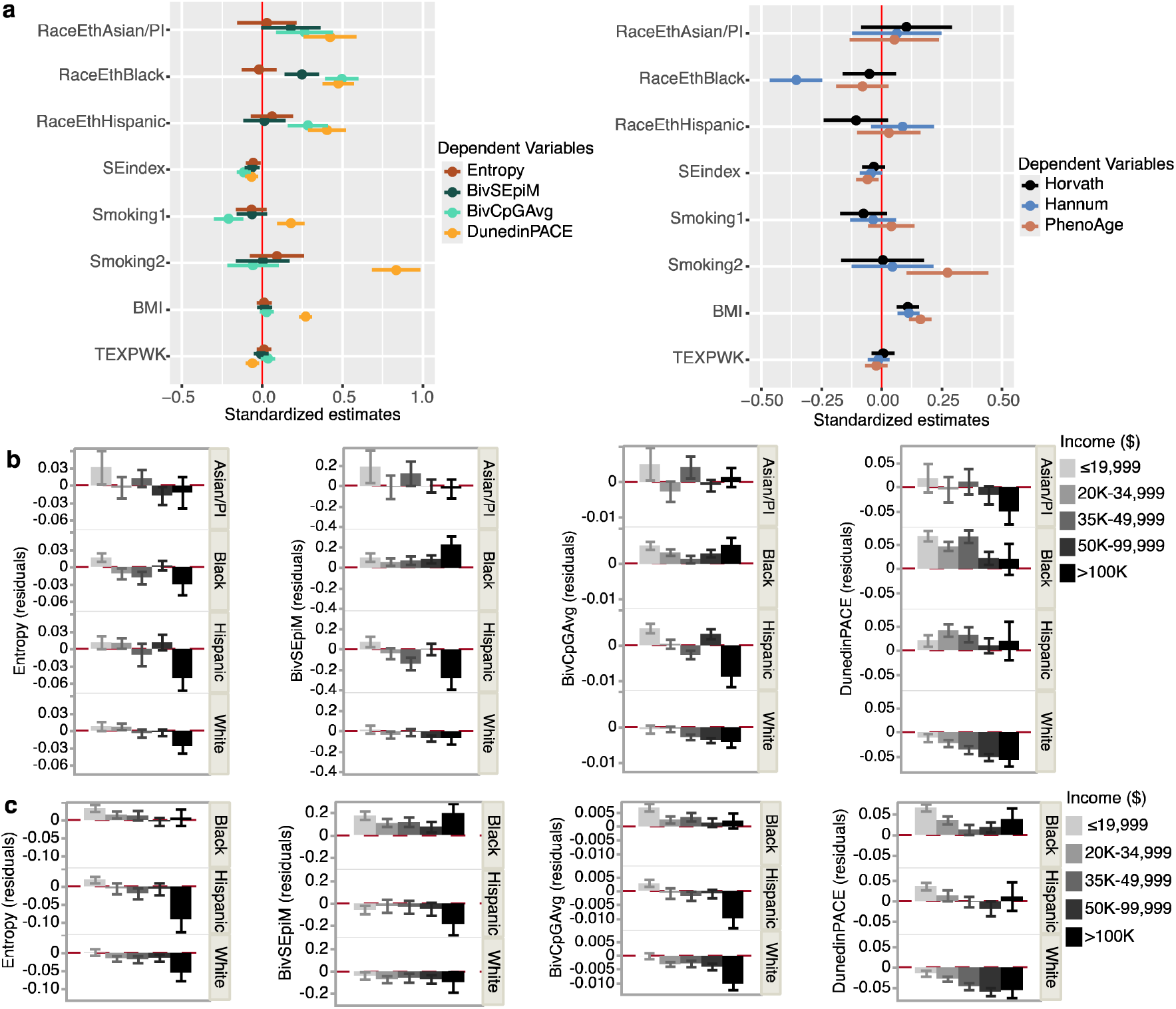
(a) Forest plots of standardized regression coefficients (95% confidence intervals) for the baseline predictor variables used in Model 1 for the AS315 group. The outcome variables are the age-adjusted DNAm readouts. Smoking1 and Smoking2 are past and present smokers, respectively, and regressions are related to never smokers. Estimates for race/ethnicity are relative to non-Hispanic White. (b) Bar graphs of ageadjusted entropy, BivSEpiM, BivAvg, and DunedinPACE plotted separately by self-reported race/ethnicity, and overlaid by income groups in AS315, and (c) in BA23 (bars shades denote income levels). Positive values indicate higher biological aging relative to chronological age. Error bars are standard errors.

**Table 3.** Association between epigenetic readouts and socioeconomic index (SE index)

| DNAm outcomes<br>(age residuals) | AS315 |  |  |  |  | BA23 |  |  |  |  |
| --- | --- | --- | --- | --- | --- | --- | --- | --- | --- | --- |
|  | Min | Max | SD | SE index<br>coef<sup>1</sup> | p | Min | Max | SD | SE index<br>coef<sup>1</sup> | p |
| DNAm Entropy | -0.655 | 0.344 | 0.10 | -0.004 | 0.018 | -0.548 | 0.452 | 0.13 | -0.005 | 0.008 |
| BivSEpiM<br>(log10 outlier<br>counts) | -1.437 | 2.312 | 0.50 | -0.02 | 0.009 | -1.453 | 2.373 | 0.51 | -0.009 | 0.216 |
| BivCpGAvg<br>(beta) | -0.032 | 0.060 | 0.01 | -0.001 | 4.20E-07 | -0.059 | 0.098 | 0.02 | -0.001 | 7.00E-07 |
| DunedinPACE | -0.358 | 0.706 | 0.13 | -0.005 | 0.002 | -0.414 | 0.485 | 0.13 | -0.009 | 4.91E-07 |
| Horvath<br>(years) | -27.71 | 93.40 | 6.22 | -0.13 | 0.16 | -29.16 | 54.18 | 6.64 | -0.11 | 0.263 |
| Hannum<br>(years) | -21.07 | 48.44 | 5.35 | -0.15 | 0.058 | -23.86 | 31.59 | 5.83 | -0.30 | 0.0007 |
| PhenoAge<br>(years) | -25.86 | 57.20 | 6.60 | -0.25 | 0.012 | -38.29 | 36.32 | 7.31 | -0.46 | 3.30E-05 |

<sup>1</sup>Unstandardized regression estimates based on Model 1

We computed these same readouts in BA23, and in the much smaller AS311. After excluding participants who were also part of the AS315, the negative associations between SE index and bivalent readouts and entropy remained consistent in both data sets **(Figs S3** and **Data S2, S3**). For example, in BA23, a unit increase in SE index resulted in 0.005 lower entropy, and 0.001 lower beta BivCpGAvg, a 0.009 slower DunedinPACE per chronological age, and a 0.46 years younger biological age relative to chronological age for the PhenoAge (**Table 3**). To illustrate the gradational effect of income on these measures of aging, we display bar plots of the non-training epigenetic readouts and the DunedinPACE (after adjusting for chronological age) by self-reported income for AS315 and BA23 (**Fig 2b, c**). The distribution and variance are shown as violin and box plots in **Fig S4**. The potential age slowing effect of high income appears particularly pronounced in the White population, whereas, for the Black participants, most of the DNAm readouts had positive age-adjusted residual values even among the highest income group (**Fig 2b**, **c**).

In Model 2, we included additional baseline health variables: history of hysterectomy, diabetes, CVD, hypertension, and cancer; and in Model 3, we included all the variables in Model 2, plus alcohol intake and the healthy eating index (2010 HEI) score to account for differences in dietary intake (**Data S1–S3; Fig S5**). These models were implemented to verify that the negative associations with SE index is robust. As sensitivity analysis, we repeated Model 2 in AS315 after excluding all participants with DNA from follow-up annual visits (**Data S1**). All these showed that the non-training based DNAm readouts, and the DunedinPACE and PhenoAge have negative associations with SE index. The entropy and bivalent readouts were not significantly associated with baseline health variables aside from slightly higher entropy for women who had undergone hysterectomy in AS315 and BA23 (but not replicated in AS311; **Fig S5**). The training-based biomarkers were more strongly related to baseline health and lifestyle variables. The DunedinPACE and PhenoAge showed the expected age-accelerating effect of smoking, and the DunedinPACE was slowed by higher total energy expenditure from recreational activity (MET-hours/week) and higher HEI score. Higher BMI was positively associated with all the training-based biomarkers, but not with entropy, BivSEpiM, or BivCpGAvg. In AS315, the BivCpGAvg showed an unexpected significant negative regression estimate for prior smokers relative to never smokers, but no difference between never and current smokers was observed; however, this was not replicated in BA23 or AS311.

### Social disparity and methylation at the chromatin states

To examine if the association with SE index was specific to the polycomb and bivalent CpGs, we performed the Models 3 regressions with the age-adjusted mean methylation values for the remaining 12 chromatin states in the AS315 and BA23 datasets. At a p threshold of 0.004 (Bonferroni corrected alpha = 0.05 for 12 tests), only the polycomb repressed state, ReprPC, had significant negative association with SE index in AS315 and this was replicated in BA23 (**Fig 3a**; **Table S1**). Notably, like the bivalent states, ReprPC is a PRC2 targeted domain, and these sites also have typically low methylation when young and accrue methylation with aging (all plot to top left quadrant of **Fig 1e**). This pattern for the bivalent and polycomb sites suggests that higher SE index is associated with a lower and presumably more “youthful” methylation state. In both AS315 and BA23, higher SE index also had negative associations with methylation levels at the active chromatin states, TssA and TssAFlnk (transcription start sites at or flanking actively transcribed genes), which are also regions with low average methylation. However, at the Bonferroni corrected alpha = 0.05, the negative association of TssA and TssAFlnk with SE index was significant only in BA23 (**Fig 3a; Table S1**).

**Fig 3.**
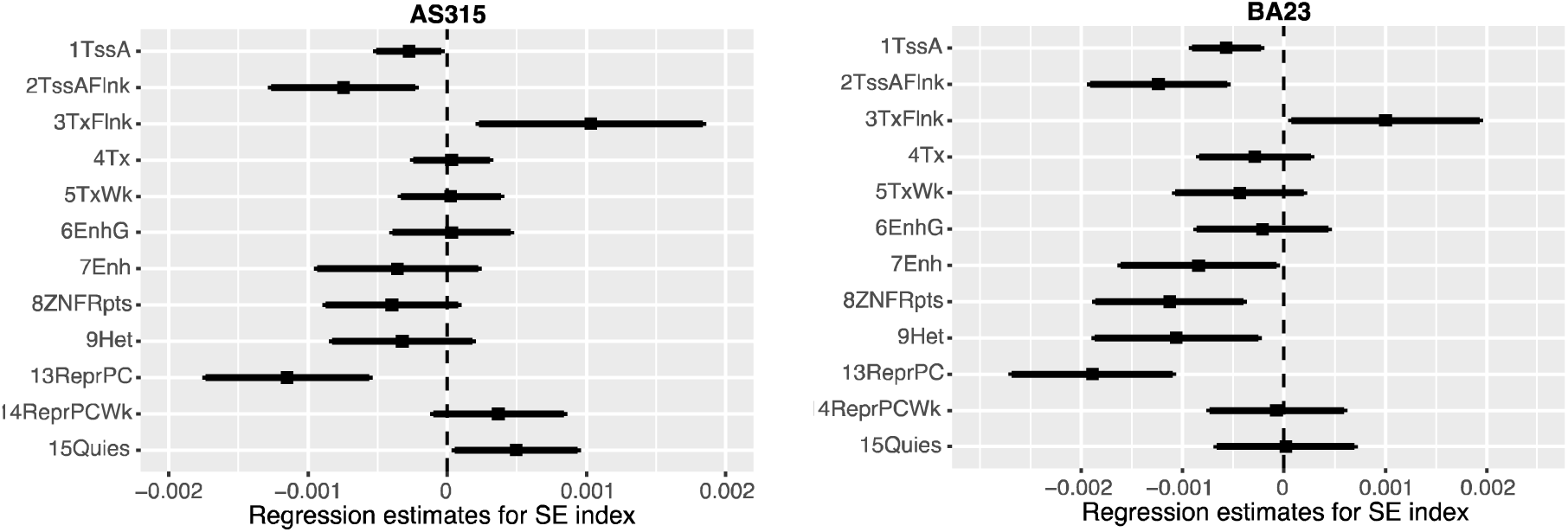
Forest plots of regression coefficients (95% confidence intervals) for SE index as predictor variable. Dependent variables are age-adjusted mean methylation at chromatin states (each fitted separately) after adjustment for other baseline variables (Model 3). One standard deviation higher SE index was associated with lower methylation at TssA, TssAFlnk, and ReprPC, and higher methylation at TxFlnk in both datasets.

Another chromatin state, TxFlnk (downstream of transcribed genes), had a positive association with SE index in both AS315 and BA23; however, the association was not significant after multiple test correction. Nonetheless, it is notable that in contrast to the bivalent states, CpGs in TxFlnk typically exist in hemi-methylated states (beta close to 0.5), loses methylation with aging, and contribute negatively to entropy (r=-0.41, **Fig 1d**). The positive association between TxFlnk and SE index suggests that higher SE index may be related to methylation maintenance at these CpGs.

### DNAm readouts and all-cause mortality

Next, we tested whether these DNAm readouts predict life expectancy. Due to the low sample numbers, we performed this analysis only for the Black, White and Hispanic participants for AS315 and BA23. The PhenoAge clock is already known to be predictive of lifespan^3,27^, and for the present work, we examined only the non-training measures, and the DunedinPACE as a training-based reference. We first performed a race/ethnicity stratified Cox regression for censored survival time, and the DNAm readouts as primary predictors (entropy, BivSEpiM, BivAvg, DunedinPACE; each fitted separately), and with SE index and baseline health variables and the Clinical Trial (CT) dietary modification and hormone replacement arms as covariates. In both AS315 and BA23, only the DunedinPACE was a significant predictor of lifespan, and one SD increase in the pace of aging increased the risk of death by about 20% with hazard ratio (HR) of 1.23 (95% confidence interval CI = 1.13–1.33) in AS315 and HR = 1.16 (1.07–1.25) in BA23 (**Fig 4a, 4b**; **Table S2**). For SE index, one SD increase was associated with a modest decreased risk of mortality of HR of 0.95 (CI = 0.87–1) in AS315, and HR 0.89 (0.83—0.96) in BA23. We further examined if SE index and DunedinPACE were associated with lifespan when race/ethnicity are analyzed separately (**Fig S6**; **Table S2**). In both AS315 and BA23, higher DunedinPACE increased risk of mortality in all groups, and the added lifespan privilege of higher income was seen mainly among the White participants.

**Fig 4.**
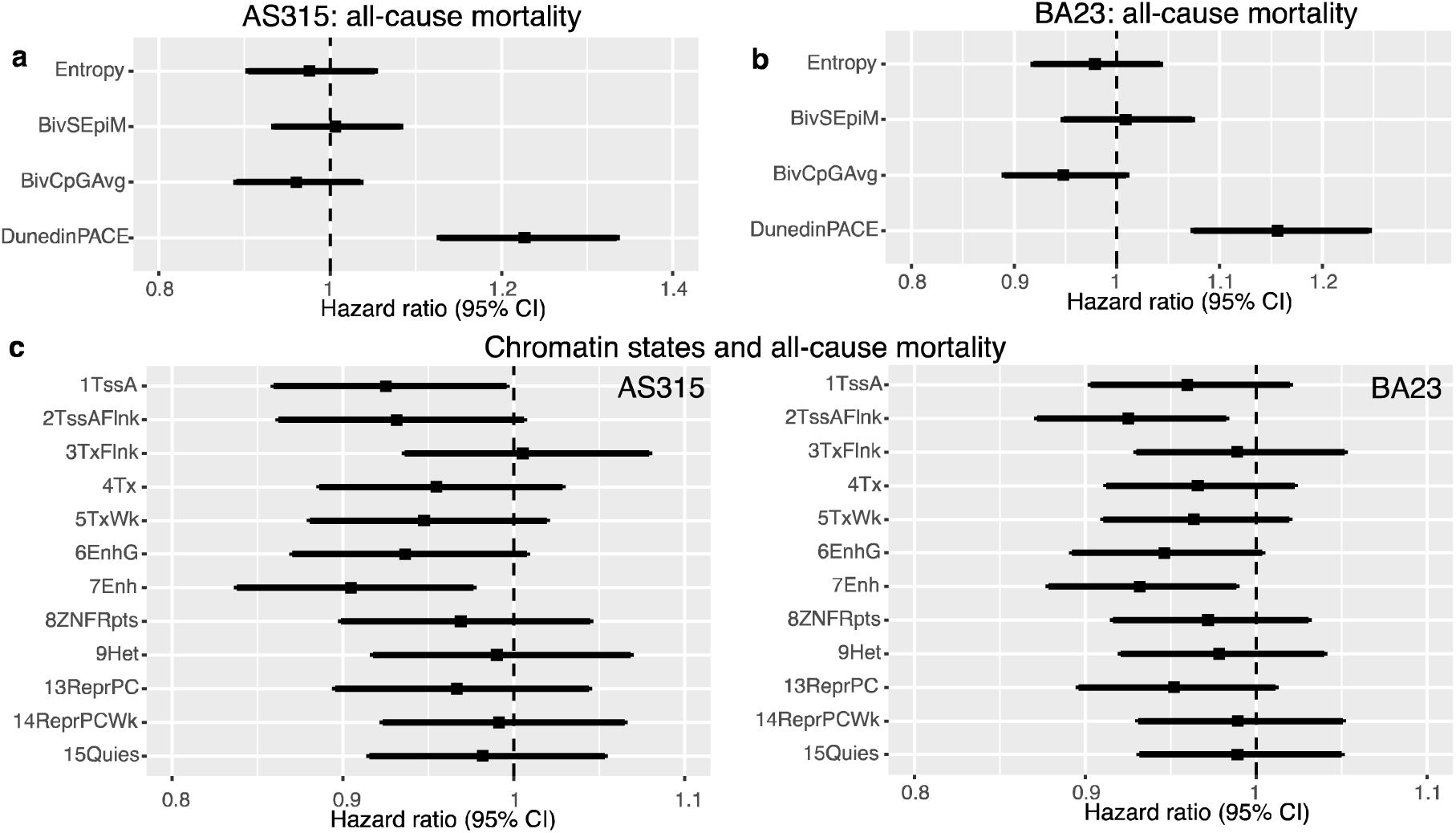
Effects of DNAm readouts on all-cause mortality risk in (a) AS315 and (b) BA23 based on race/ethnicity stratified multivariable Cox regression. The predictor variables (entropy, BivSEpiM, BivCpGAvg, and DunedinPACE) were fitted separately and the forest plots displays the hazard ratios associated with each. Only the DunedinPACE is predictive of survival time in both datasets. (c) HR associated with mean methylation at chromatin states. Higher methylation at Enh (enhancers) has a nominally significant association with reduced risk of death in both AS315 and BA23.

We applied the same Cox regression to examine whether the average methylation levels of the 12 remaining chromatin states are predictive of all-cause mortality. Generally, higher methylation at chromatin states that were negatively correlated with chronological age were associated with reduced mortality risk (**Table S3**). This was nominally significant in both AS315 and BA23 for enhancer (Enh) CpGs (**Fig 4c**; **Table S3**). Higher mean methylation of Enh states predicted lower mortality risk with HR of 0.90 (0.84—0.98) in AS315, and HR of 0.93 (0.88—0.99) in BA23. Notably, the mean methylation of Enh regions also had the strongest negative correlation with chronological age **(Fig 1f**). While the association with mortality risk is not significant after Bonferroni correction, the overall pattern suggests that maintaining higher methylation at these regulatory CpGs that typically lose methylation with aging could be related to longer life expectancy.

## DISCUSSION

To summarize: **(1)** We found that nearly all the large-scale methylation patterns (i.e., entropy, overall methylation averages of chromatin states, and outlier counts at the bivalent sites) showed some degree of change with chronological age among postmenopausal women and the typical pattern was for the low methylation CpGs to gain, and high methylation CpGs to lose methylation with aging. **(2)** Age-dependent changes in the bivalent and polycomb targeted sites contributed to the increase in epigenetic entropy. **(3)** The bivalent and polycomb CpGs were consistently associated with SE index and varied between self-reported race/ethnicity. **(4)** However, the untrained readouts were not significantly associated with baseline health variables or predictive of mortality risk. **(5)** In contrast, biomarkers trained on health-related parameters (i.e., the DunedinPACE and PhenoAge) were significantly associated with baseline variables such as BMI, smoking, and energy expenditure; furthermore, a higher pace of aging estimated by the DunedinPACE was associated with a higher risk for all-cause mortality. **(6)** Among the chromatin states, we found evidence that maintaining methylation levels at enhancer CpGs could be associated with a lower risk of death.

Of the DNAm readouts, the most “global” measurement we computed was the methylome-wide entropy that was defined using nearly the full set of CpGs. This value estimates the overall randomness and is related to the phenomenon of “epigenetic drift”, the increase in stochasticity and variance in DNAm with aging.^14^ In cohorts with a wide age range, epigenetic entropy has a strong positive correlation with chronological age.^1^ Among the WHI participants, the positive correlation between entropy and age was modest, albeit significant. This is possibly because the WHI represents an older cohort, and the relationship between epigenetic aging and chronological age is not perfectly linear and there is evidence that it begins to plateau among older individuals.^9,43^ Initially we had anticipated that health variables such as BMI and history of cancer would show some association with epigenetic entropy. However, this was not the case. Instead, the combination of self-reported education and income (what we refer to as SE index) was the only variable at baseline that had a consistent inverse association with global entropy in all three WHI datasets. Scoring high on the SE index was linked to lower age-adjusted entropy. Since the increase in DNAm entropy will partly reflect the increase in cellular heterogeneity with aging, we also attempted to account for blood cell heterogeneity by including the DNAm-based estimated proportions of blood cell types, and in doing so, the link between epigenetic entropy and SE index became stronger (see **Data S2–S3**). The lowest age-adjusted entropy was seen among the White and Hispanic women at the highest income grade, while the highest age-adjusted entropy was seen among the low-income minority groups. This suggests that while higher income and the numerous psychosocial, environmental, and health privileges of wealth could be protective against the buildup of epigenetic discordance with aging, the impact of income is not the same across all racial groups. In fact, the effect is blunted especially among Black women. This is interesting, yet not surprising as Black women who achieve high education and socioeconomic status continue to experience chronic stressors over their lifetime due systemic racism, discrimination, microaggressions and cultural expectations.^44,45^

SE index also had a similar inverse relationship with methylation levels at bivalent CpGs, which were the strongest positive correlates of methylome-wide entropy. Having a low SE index was characterized by higher levels of stochastic epimutations and age-dependent gains at these bivalent sites. Here, stochastic epimutation simply refers to the counts of outlier CpGs and this is another way of quantifying the levels of epigenetic discordance.^40,41^ Our finding is consistent with previous work by Fiorito et al. ^40^ Theirs was a multi-cohort study that included both males and females, and participants from a wide age-range. They also used education level as a proxy of socioeconomic position, and found that methylome-wide stochastic epimutation was higher among those with lower education.^40^

We must note that both education and income are readily recordable variables that are part of an extremely complex social construct of ones status in society and referred to as socioeconomic status.^46^ The SE index therefore serves only as a global proxy that tags along with a multitude of unmeasured variables such as differences in access to health care and education, food and nutrition options, neighborhood and housing conditions, environmental pollutants and toxins, chronic stress, etc. These exposures—the social and environmental determinants of health—exert a strong influence on interrelated biological processes such as the stress and glucocorticoid pathway, composition of circulating immune cells and inflammatory state, metabolic health, etc. ^47–49^ Our results suggest that these large-scale non-training based readouts from the epigenome are influenced by the larger socio-environmental context, but lack the sensitivity to serve as predictors of specific health and lifestyle conditions such as BMI, energy expenditure, smoking, and disease status.

In contrast to the global entropy and bivalent readouts, the widely used epigenetic clocks and pace-of-aging biomarkers are derived from extensive training algorithms. For the initial training, both the PhenoAge and DunedinPACE included blood biomarkers that are related to health and correlated with aging (e.g., C-reactive protein, blood cell counts, albumin, cholesterol).^3,4,50^ The DunedinPACE also included waist-hip ratio, dental health, lung function, cardiorespiratory fitness, etc. Notably, the DunedinPACE was trained on the New Zealand (NZ) based Dunedin Longitudinal Cohort, which consists predominantly of European ancestry participants, with only a very small minority of participants identifying as indigenous people of New Zealand.^4,50^ It is quite remarkable that a biomarker panel of aging developed on a NZ cohort can generalized to the societal structure and inequities in the United States. The DunedinPACE also revealed a potential “anti-aging” effect of higher income among the White women, but the effect of income was not as pronounced among the minority women. Except for the small group of self-identified Asians on the higher income scale, all other minority groups had positive pace of aging according to the DunedinPACE. While the training population could partly contribute to the differences between race/ethnic groups, there is evidence that DunedinPACE captures the effects of adversity and negative experiences that is independent of genetic ancestry.^51^ Having a higher pace of aging likely has important health consequences, and in both AS315 and BA23, higher pace of aging as measured by DunedinPACE predicted shorter lifespan independent of SE index.

Segregating the CpGs by their predicted chromatin states and computing the average methylation levels was a very broad stroke approach. However, we applied this data reduction to determine the general patterns by which chromatin states contribute to the increase in epigenetic entropy. The strongest positive correlates of entropy were the methylation levels at the bivalent chromatin domains and regions bound by the PRC2 (TssBiv, BivFlnk, EnhBiv, ReprPC). These sites mark repressed states with low average methylation, and have an age-dependent drift towards higher methylation. The bivalent and polycomb target sites are highly conserved regulatory regions, and are crucially involved in embryonic development and in maintaining cellular identity and function.^13^ Similar to the bivalent and polycomb domains, CpGs at active promoters (TssA) also typically have low methylation levels, and in the WHI, the mean methylation at TssA was positively correlated with entropy. Notably, the chromatin states that had significant positive correlations with entropy (i.e., higher methylation related to higher entropy) were all negatively associated with SE index. An interpretation we draw from this is that higher SE index is associated with the age-dependent increase in discordance and drift at these CpG sites. In contrast, the chromatin state with the strongest negative correlation with entropy was TxFlnk, and this was the only chromatin domain that showed a significant and replicated positive association with SE index (means higher SE index related to higher methylation levels at TxFlnk). TxFlnk marks CpGs located at 3’ and 5’ ends of transcribed genes, and the mean methylation of TxFlnk had a weak inverse correlation with age (i.e., methylation loss with aging), and maintaining a higher methylation reduced the level of entropy. The positive association with SE index suggests that high SE index could help maintain a higher methylation level at these CpGs, and thereby contribute to lower entropy.

Intriguingly, when it came to predicting survival time, the general trend was for higher methylation at active chromatin states (e.g., TssA, TssAFlnk, EnhG, and Enh) to be linked to lower risk of all-cause mortality, and this reached statistical significance for Enh. Except for TssA, which was uncorrelated with chronological age, all these chromatin states tended to have a negative correlation with aging, and this was strongest for Enh. Enhancers are important regulatory elements, and the ENCODE 15-states model divides them into three categories based on histone marks: bivalent enhancers in repressed sites (EnhBiv) that have low methylation, and enhancers in genic (EnhG) and non-genic (Enh) sites.^35^ Both Enh and EnhG mark actively transcribed regions, and other studies have also shown that these have higher methylation and lose methylation with aging.^52–54^ Taken together, our results suggest that being able to maintain higher methylation levels at these enhancer CpGs may extend lifespan. To our knowledge, this is the first time the global average methylation of Enh CpGs has been associated with all-cause mortality in humans. There is however evidence from mouse studies that interventions such as caloric restriction, which extends lifespan, suppress the age associated methylation loss at enhancer CpGs.^52^ Furthermore, in humans, aberrant methylation at enhancer CpGs have been implicated in cancer progression and metastasis.^55^ The work by Cole et al.^55^ also showed that higher methylation at the enhancer CpG of the metastasis gene *KIT* increased survival time among cancer patients. This direction of association is therefore in agreement with our observation that higher methylation at Enh is associated with a decreased risk of all-cause mortality.

In the present work, we only examined time to all-cause mortality but did not perform a more detailed analysis to test whether these DNAm readouts predict time to adjudicated diseases or disease-related deaths. Given the links between enhancer methylation and cancer progression and mortality, this will be particularly relevant in a follow-up analyses. We also acknowledge that the results we present are all based on older postmenopausal women (age 50-79 at baseline), and the significant associations between the bivalent and polycomb sites and SE index may represent the effects of lifelong exposures to social inequities and weathering. Further work is needed to examine whether the link between SE index and the bivalent CpGs is also seen in younger populations. For the DunedinPACE, there is strong and growing evidence that it serves as a sensitive biomarker of the age-accelerating effects of social inequities and discrimination in a wide age range, and in both males and females.^28,29,56–58^ Based on this corpus of work, we also anticipate that the association between the social variables and the non-training based DNAm readouts present in this work will also be generalizable, but that is yet to be tested. The link between enhancers and lifespan also needs further replication. To facilitate such follow-up studies, we have provided all the computational codes, and these can be easily computed from existing datasets and can be adapted to the newer DNAm microarray datasets.

In conclusion, we have presented alternative ways to quantify aspects of epigenetic aging that are based on epigenetic entropy and stochasticity, and have related the large-scale methylation features to aging, as well as to extrinsic variables, and potentially to lifespan. The presented work highlights the deep links between the larger social environment and the aging of the epigenome and provides evidence that methylation at active chromatin states could be related to life expectancy.

## METHODS

### WHI DNA methylation datasets and sample inclusion

The WHI is a multicenter long-term prospective study of postmenopausal women that began in 1993.^18–20^ Women between the ages 50–79 were recruited in 1993–1998. Ancillary studies have generated genomic data, and the present work uses the genome-wide DNA methylation dataset by the three ancillary studies : (1) WHI-EMPC/AS315,^26,31^ (2) BA23,^21,32^ and (3) AS311. These 3 datasets have been previously analyzed jointly as part of large meta-studies, and detailed descriptions can be found in these publications.^3,59–61^

The EMPC study (Epigenetic Mechanisms of PM-mediated CVD Risk; AS315) randomly selected 2200 participants from within the Clinical Trial arm and measured DNA methylation at SV for the majority or at annual visit 3 or 6, and with repeated measurements for a subset of the samples ^31^. We used only the first sampling for the present work, and after excluding eight samples with low methylation calls, the primary analyses were done for 2192 AS315 participants. The statistical analysis for AS315 was repeated in the 1396 SV samples (**Fig S1**). BA23 was designed as a case/control study of risk for future CHD. We received epigenetic data for 2107 BA23 participants collected at SV, and after excluding samples that overlap with WHI-AS315, we performed the analyses in 1989 participants. AS311 is a matched case-control study of bladder cancer, and like BA23, used blood DNA collected at SV and prior to disease diagnosis. We received 882 array data; after excluding an array with too many missing values (∼262,970 CpGs had NA values) and samples that overlap with AS315, we performed analyses in 868 participants.

### Computing entropy, chromatin annotations, and estimates of biological aging

All analyses were done using the processed data that were provided in NetCDF format. For uniformity, we used the beta-mixture quantile (BMIQ) normalized data, which applied the same data quality control and processing steps implemented by WHI-EMPC ^31^. The R package “ncdf4” was used to interface with the data matrices ^62^. Before any downstream analyses, we first counted how many CpGs had missing values per sample, and excluded one sample from AS311, and eight samples from BA23 as those had unusually high missingness (>50%).

For methylome-wide discretized entropy calculation, we used the same method described in a prior study.^16^ The optimal number of bins was estimated to be 50 using the Freedman-Diaconis rule ^63^. The R codes used for each step are in supplemental file **DataS5**. We then used the R package “entropy” ^64,65^ to compute the discretized entropy value for each participant and this was scaled to a number between 0–1. Example density plots for the beta-values were generated in R with ggplot2 ^66^ for ten participants of AS315; these were all non-Hispanic Whites and five had low entropy, and five had high entropy.

To annotate for chromatin states, we downloaded the archived REMCChromHMM file from the Zhou github page ^38,67^. This contains the 15-states chromHMM consensus chromatin annotations for each of the ∼480,000 CpGs in the 450K platform ^35^. These annotations were matched by CpG probe IDs to the WHI BMIQ data matrices, and average methylation levels for each chromatin state was computed for each participant. As an alternate measure of randomness in the epigenetic data, we counted CpGs that were outliers in the three bivalent states to compute epigenetic stochasticity: bivalent/poised transcription start site (TssBiv), flanking bivalent TSS (BivFlnk), and bivalent enhancer (EnhBiv). While the entropy measures the shift in the genome-wide methylome landscape for each participant and is not relative to the study cohort, the outlier counts are estimates of how many CpGs are outliers for an individual relative to the study cohort. We adapted the outlier detection method described in Gentilini but counted only the positive outliers as these sites gain methylation with aging and a higher beta-value is presumed to represent a more aged methylome, i.e., beta-values over 3-times the interquartile range of the 3^rd^ quartile ^40,41^. The codes used to compute the bivalent stochastic epimutations (BivSEpiM) are in **Data S4**.

For the first-generation epigenetic clocks (i.e., Horvath and Hannum), we used the pre-computed clocks that were shared by the WHI. Specifically, we used the linear transformed Horvath and Hannum DNAm age predictions ^1,2^. We computed the PhenoAge (aka, Levine2018) and the DunedinPACE using the dnaMethyAge R package ^3,4,68,69^.

### Statistical analyses

Pearson correlations were used for bivariate comparisons of chronological age with the DNAm readouts. The pair-wise correlation plots were generated using the corrplot R package ^70^, and display significance threshold was set at p=0.001. For epigenetic clocks, which correlate strongly with chronological age, the deviation of the predicted biological age from chronological age (referred to as age-acceleration or age-deviation) is computed as the residuals of predicted age regressed on chronological age ^2^. As most of the DNAm readouts, and the DunedinPACE also show varying levels of correlation with chronological age, we used the residuals to make the values independent of chronological age.

We used the linear regression lm() function in R to test associations between the age-residuals of the DNAm readouts and baseline variables. These baseline variables included BMI computed from weight and height at baseline (kg/m^2^), medical history at baseline, and personal habits including smoking status (0 = never smoked; 1 = past smoker; and 2 = present smoker), and recreational energy expenditure computed as measured by the TEXPWK variable (MET-hours/week). TEXPWK was computed from questionnaire-based reports of recreational physical activity that included walking, mild, moderate and strenuous physical activity in kcal/week/kg.^71^ Among the demographic variables, WHI collected self-reported education and income at time of screening and are presented on an ascending scale from 1–11 for education, and 1–8 for income (higher numbers represent higher education or income). We derived a single variable, SEindex as the average of the two after excluding unreported or missing data.

We applied a simple model, Model 1: lm(y_i_ ∼ race/ethnicity + SEindex + Smoking + BMI + TEXPWK), and y_i_ is each of the DNAm readouts fitted independently. We limited the analysis to participants who identified as non-Hispanic White (White), non-Hispanic Black (Black), Hispanic, and Asian or Pacific Islander (Asian/PI) in AS315, and White, Black and Hispanic for BA23 and AS311. Model 2 was the same as Model 1 and included additional medical history terms collected at baseline: hysterectomy, diabetes, cardiovascular disease (CVD), hypertension, and cancer ever. An expanded version of Model 1 included blood cell proportions (lymphocytes, monocytes, granulocytes) that were indirectly estimated from the DNAm data. The DNAm data for BA23 and AS311 were from baseline SV, and majority of the AS315 blood DNA was from the SV prior to assignment of a study arm (n=1396). Model 2 was repeated for AS315 after excluding the 633 blood DNA that were sampled at visit year 3, and 163 at visit year 4. Model 3 included all the variables in Model 2, plus the alcohol intake variable, and 2010 HEI total score.^72,73^ The lm() R codes and details on the variables are provided in **Data S4**.

We performed Cox regressions to test predictive value of the DNAm readouts using the 2022 versions of All Discovered Deaths, and Adjudicated Outcomes. This was done using the survival R package ^74,75^. For all-cause mortality, we used the variable DEATHALL (all discovered deaths including from National Death Index) in conjunction with ENDFOLLOWALLDY (days enrollment to end of follow up including uncensored death). Covariates for Cox regression were baseline variables age, smoking status, BMI, diabetes, cancer, SE index, hormone therapy arm, dietary modification arm, alcohol intake, 2010 HEI total score, and with race/ethnicity in the strata() term. The coxph() R codes are provided in **DataS4**.

## Data availability

All data are available upon request through the Women’s Health Initiative Study (https://www.whi.org).

## Supporting information

Supplemental Data S4 (R codes)

## Acknowledgement

The WHI program is funded by the National Heart, Lung, and Blood Institute, National Institutes of Health, U.S. Department of Health and Human Services through contracts HHSN268201600018C, HHSN268201600001C, HHSN268201600002C, HHSN268201600003C, and HHSN268201600004C. WHI EMPC (AS315) was supported by NIEHS grant R01-ES020836 (Whitsel, Eric A and other PIs). WHI AS311 was supported by American Cancer Society award 125299-RSG-13–100-01-CCE (Parveen Bhatti). WHI-BAA23 was supported by NHLBI Broad Agency Announcement contract HHSN268201300006C.

## Author contribution

KM: study design, analyses and interpretation, initial manuscript preparation, review and editing. AD, YS, AHS, RC, FJT, RBW, JHF: part of WHI writing group and contributed to manuscript reviewing, editing, and interpretation. KCJ: WHI sponsor and contributed to manuscript reviewing, editing, and interpretation. All authors approved final manuscript.

