## Supplemental Figures S1 to S6, and Tables S1 to S3 for "Epigenetic entropy, social disparity, and health and lifespan in the Women’s Health Initiative"

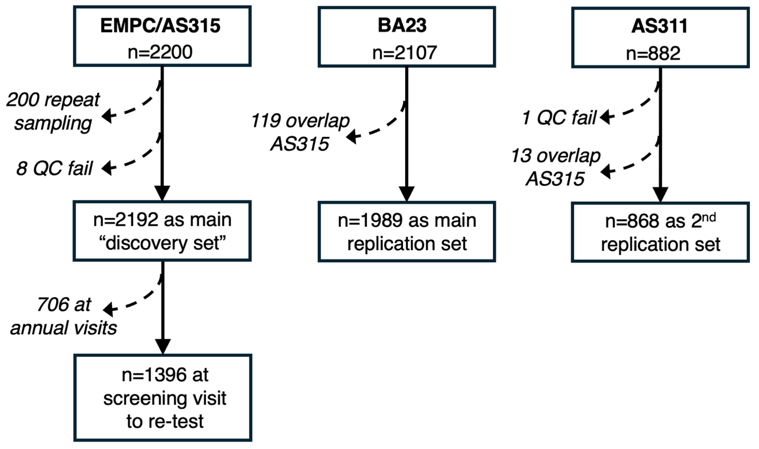

**Fig S1.** Sample numbers in the three datasets and inclusion/exclusion.

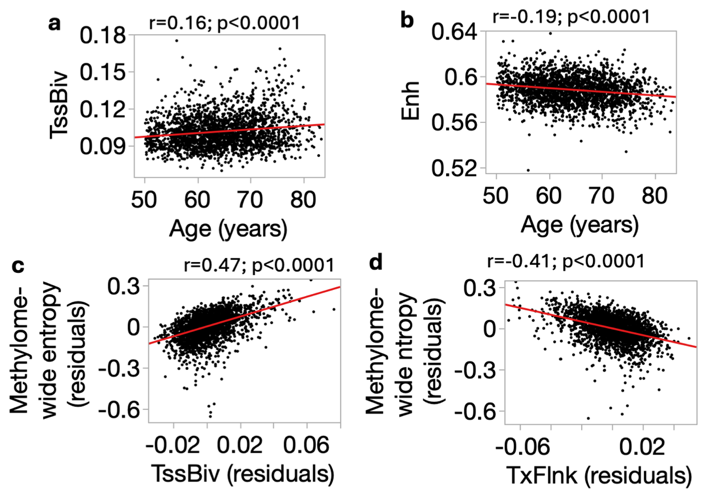

**Fig S2.** Correlations between chronological age and mean methylation at **(a)** TssBiv, and **(b)** Enhancers. Correlations between age-adjusted entropy and age-adjusted mean methylation at **(c)** TssBiv and **(d)** TxFlnk. These are from AS315.

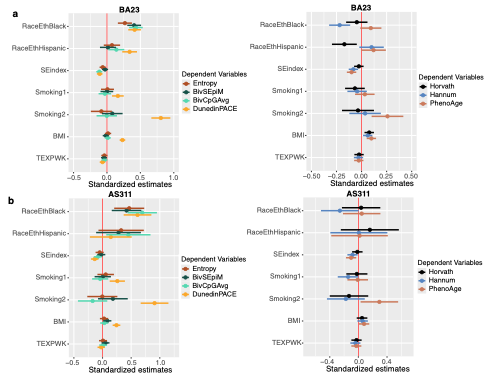

**Fig S3.** Forest plots of standardized regression coefficients (95% confidence intervals) for the baseline predictor variables used in Model 1 for the **(a)** BA23 and **(b)** AS311. The outcome variables are the age-adjusted DNAm readouts. Smoking1 and Smoking2 are past and present smokers, respectively, and regressions are related to never smokers. Estimates for race/ethnicity are relative to non-Hispanic White.

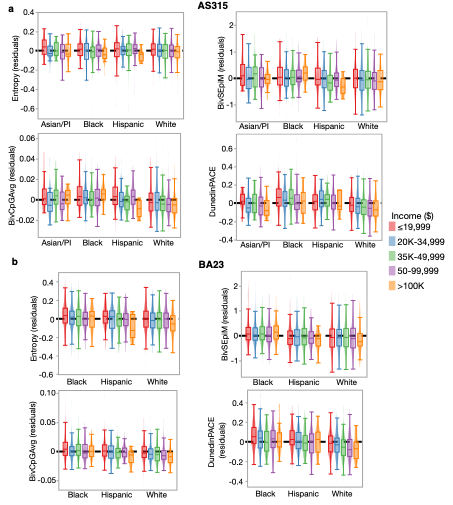

**Fig S4.** Violin and bar plots of the age adjusted epigenetic readouts: entropy, BivSEpiM, BivAvg, and DunedinPACE in **(a)** BA23 and **(b)** AS311. Plotted separately by race/ethnicity and overlaid by self-reported income levels.

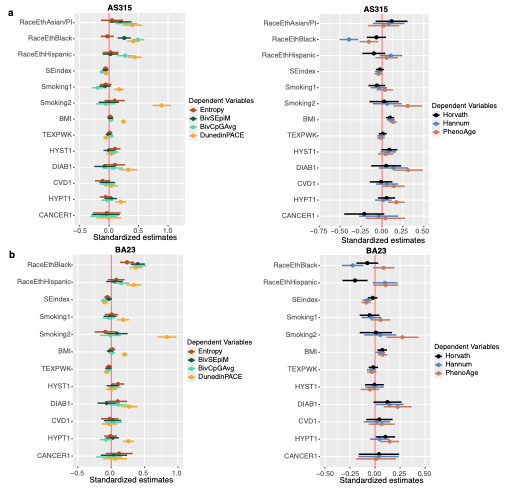

**Fig S5.** Forest plots of standardized regression coefficients (95% confidence intervals) for the baseline predictor variables used in Model 2 for the **(a)** AS315 and **(b)** BA23. The outcome variables are the age-adjusted DNAm readouts. Smoking1 and Smoking2 are past and present smokers, respectively, and regressions are related to never smokers. Estimates for race/ethnicity are relative to non-Hispanic White. Baseline health variables are: HYST1 is Hysterectomy “Yes” DIAB1 is diabetes “Yes”; CVD is cardiovascular disease “Yes”; HYPT1 is hypertension “Yes”, and CANCER1 is cancer “Yes” relative to “No”.

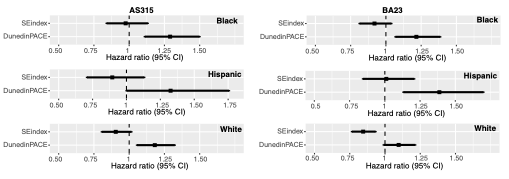

**Fig S6.** Effects of SE index and DunedinPACE on all-cause mortality based on multivariable Cox regression that adjusted for baseline variables and applied separately in the difference race/ethnicity groups.

**Supplemental tables**

**Table S1. Association between mean methylation of chromatin states and socioeconomic index (SE index)**

| **ChromHMM** | **AS315** | | | **BA23** | | |
| --- | --- | --- | --- | --- | --- | --- |
|  | **Coef^1^** | **Std. Error** | **p^2^** | **Coef^1^** | **Std. Error** | **p^2^** |
| TssA | -1.73E-04 | 7.5E-05 | 0.02 | -3.60E-04 | 1.10E-04 | 0.001 |
| TssAFlnk | -4.72E-04 | 1.7E-04 | 0.005 | -7.86E-04 | 2.18E-04 | 0.0003 |
| TxFlnk | 6.54E-04 | 2.6E-04 | 0.01 | 6.36E-04 | 3.01E-04 | 0.04 |
| Tx | 2.01E-05 | 8.9E-05 | 0.82 | -1.82E-04 | 1.78E-04 | 0.308 |
| TxWk | 1.70E-05 | 1.2E-04 | 0.88 | -2.76E-04 | 2.05E-04 | 0.179 |
| EnhG | 2.03E-05 | 1.4E-04 | 0.88 | -1.33E-04 | 2.11E-04 | 0.529 |
| Enh | -2.26E-04 | 1.9E-04 | 0.23 | -5.34E-04 | 2.49E-04 | 0.032 |
| ZNFRpts | -2.51E-04 | 1.5E-04 | 0.10 | -7.15E-04 | 2.36E-04 | 0.002 |
| Het | -2.04E-04 | 1.6E-04 | 0.21 | -6.73E-04 | 2.61E-04 | 0.01 |
| ReprPC | -7.28E-04 | 1.9E-04 | 0.0001 | -1.20E-03 | 2.56E-04 | 0.000003 |
| ReprPCWk | 2.33E-04 | 1.5E-04 | 0.124 | -4.60E-05 | 2.14E-04 | 0.830 |
| Quies | 3.13E-04 | 1.4E-04 | 0.028 | 1.15E-05 | 2.19E-04 | 0.958 |

^1^Unstandardized regression estimates based on Model 3: *lm(ChromHMM ~ SEindex + RaceEth + Smoking + BMI + TESPWK + HYST + DIAB + CVD + HYPT + ALCOHOL + HEI2010_TOTAL_SCORE*)

^2^Nominal p-values; p=0.004 corresponds to Bonferroni corrected alpha=0.05

**Table S2. Associations between SE index and DunedinPACE, and censored survival time**

| **Predictor** | **AS315** | | | **BA23** | | |
| --- | --- | --- | --- | --- | --- | --- |
|  | **Hazard Ratio** | **p** | **Confidence interval** | **Hazard Ration** | **p** | **Confidence Interval** |
|  | Race/ethnicity stratified^1^ | | | | | |
| SE index | 0.95 | 0.19 | 0.87, 1.03 | 0.89 | 0.001 | 0.83, 0.96 |
| DunedinPACE | 1.23 | 1.82E-06 | 1.13, 1.33 | 1.16 | 0.0001 | 1.07, 1.25 |
|  | Blacks only^2^ | | | | | |
| SE index | 0.97 | 0.74 | 0.84, 1.13 | 0.92 | 0.18 | 0.81, 1.04 |
| DunedinPACE | 1.29 | 0.0009 | 1.11, 1.5 | 1.22 | 0.003 | 1.07, 1.39 |
|  | Non-Hispanic Whites only^2^ | | | | | |
| SE index | 0.90 | 0.09 | 0.81, 1.02 | 0.85 | 0.0009 | 0.77, 0.93 |
| DunedinPACE | 1.18 | 0.004 | 1.05, 1.32 | 1.10 | 0.077 | 0.99, 1.22 |
|  | Hispanics only^2^ | | | | | |
| SE index | 0.90 | 0.36 | 0.71, 1.13 | 1.01 | 0.91 | 0.84, 1.21 |
| DunedinPACE | 1.33 | 0.05 | 1, 1.76 | 1.39 | 0.002 | 1.13, 1.7 |

^1^coxph(Surv(ENDFOLLOWALLDY, DEATHALL) ~ scale(Seindex) + scale(DunedinPACE) + Age + Smoking + BMI + DIAB + CVD + CANCER + HRTARM + DMARM + ALCOHOL + HEI2010_TOTAL_SCORE + strata(Race/Ethnicity))

^2^coxph(Surv(ENDFOLLOWALLDY, DEATHALL) ~ scale(Seindex) + scale(DunedinPACE) + Age + Smoking + BMI + DIAB + CVD + CANCER + HRTARM + DMARM + ALCOHOL + HEI2010_TOTAL_SCORE, subset(Race/Ethnicity =="eth"), where "eth" is either White, Black, or Hispanic

**Table S3. Associations between mean methylation at chromatin states and censored survival time**

| **Predictor** | **AS315** | | | **BA23** | | |
| --- | --- | --- | --- | --- | --- | --- |
|  | **Hazard Ratio^1^** | **p** | **CI** | **Hazard Ratio^1^** | **p** | **CI** |
| TssA | 0.93 | 0.04 | 0.86, 1 | 0.96 | 0.18 | 0.9, 1.02 |
| TssAFlnk | 0.93 | 0.07 | 0.86, 1.01 | 0.93 | 0.01 | 0.87, 0.98 |
| TxFlnk | 1.01 | 0.88 | 0.94, 1.08 | 0.99 | 0.72 | 0.93, 1.05 |
| Tx | 0.95 | 0.22 | 0.89, 1.03 | 0.97 | 0.23 | 0.91, 1.02 |
| TxWk | 0.95 | 0.15 | 0.88, 1.02 | 0.96 | 0.20 | 0.91, 1.02 |
| EnhG | 0.94 | 0.08 | 0.87, 1.01 | 0.95 | 0.07 | 0.89, 1 |
| Enh | 0.90 | 0.01 | 0.84, 0.98 | 0.93 | 0.02 | 0.88, 0.99 |
| ZNFRpts | 0.97 | 0.41 | 0.9, 1.04 | 0.97 | 0.34 | 0.92, 1.03 |
| Het | 0.99 | 0.80 | 0.92, 1.07 | 0.98 | 0.48 | 0.92, 1.04 |
| ReprPC | 0.97 | 0.39 | 0.9, 1.04 | 0.95 | 0.11 | 0.9, 1.01 |
| ReprPCWk | 0.99 | 0.81 | 0.92, 1.06 | 0.99 | 0.72 | 0.93, 1.05 |
| Quies | 0.98 | 0.61 | 0.92, 1.05 | 0.99 | 0.72 | 0.93, 1.05 |

^1^coxph(Surv(ENDFOLLOWALLDY, DEATHALL) ~ scale(Seindex) + scale(mean methylation) + Age + Smoking + BMI + DIAB + CVD + CANCER + HRTARM + DMARM + ALCOHOL + HEI2010_TOTAL_SCORE + strata(Race/Ethnicity))
