## Supplemental Data S4 (R codes) for "Epigenetic entropy, social disparity, and health and lifespan in the Women’s Health Initiative"

###########################################################
###PART 1: Steps for calculating methylome-wide entropy####
##These were applied in AS315, BA23, and AS311 seperately##
###########################################################


#Step1: estimate optimal bin number#
betameans <- data.frame(rowMeans(tbeta)) #tbeta is the transposed BMIQ file where CpGs are rows and participants/samples are columns#
hisbk <- hist(betameans[,1], breaks="fd") #make histogrm for the population-wide mean CpG values determine number of bins##
length(hisbk$breaks)


##Step 2: use 50 bins, and compute entropy values for each participant, and scale values to 0-1##
library("entropy")
library("scales")

tbeta <- na.omit(tbeta) #this step must be done after excluding any participant/sample that has too many missing CpGs; otherwise this will remove too many CpGs##
SampleEntr <- data.frame(apply(tbeta, 2, function(x) entropy(discretize(x, numBins=50)))) ##for each participant, compute methylome-wide entropy##
SampleEntr <- data.frame(rescale(SampleEntr[,1], to=c(0,1)))


###########################################################
###PART 2: Steps for calculating stochastic epimutations###
##These were applied in AS315, BA23, and AS311 seperately##
###########################################################

#Step1: get CpGs that are in the three bivalent chromatin states## 
tbetaBiv <- t(beta[,colnames(beta) %in% rownames(bivChromhmm3)]) #beta is the BMIQ file where CpGs are columns, participants are rows; bivChromhmm3 is a dataframe with CpG probe IDs in column 1, and corresponding ChromHMM annotation is column 2#

tbetaBiv <- na.omit(tbetaBiv) #this step must be done after excluding any participant/sample that has too many missing CpGs##


#Step2: for each CpG, compute the number that is the 3rd quartile + 3-time the interquartile range#
upper3 <- data.frame("upper" = apply(tbetaBiv, 1, function(x) quantile(x,0.75) + 3*IQR(x)))


#Step3: convert the subsetted transposed BMIQ beta matrix for the bivalent sites into a TRUE=1 or FALSE=0 matrix#
df1 <- (tbetaBiv[] > upper3[,])*1


#Step4: for each participant, count the number of CpG that were outliers#
BivSEpiM <- data.frame("outlierCounts"=apply(df2,2, function(x) sum(x))) 
##counts were log10 transformed prior to downstream analyses##


###########################################################
#PART 3: basline variables as predictors of DNAm readouts##
##These were applied in AS315, BA23, and AS311 seperately##
###########################################################

#Model 1:
lm(scale(y) ~ RaceEth + scale(SEindex) + Smoking + scale(BMI) + scale(TEXPWK)) #standardized regression for graphing#
lm(y ~ RaceEth + SEindex + Smoking + BMI + TEXPWK) #for unscaled regression reports#


#Model 1B:
lm(y ~ RaceEth + SEindex + Smoking + BMI + TEXPWK + LYMPHO + MONO + GRAN) #for unscaled regression reports#


#Model 2:
lm(scale(y) ~ RaceEth + scale(SEindex) + Smoking + scale(BMI) + scale(TEXPWK) + HYST + DIAB + CVD + HYPT + CANCER) #standardized regression for graphing#
lm(y ~ RaceEth + SEindex + Smoking + BMI + TEXPWK + HYST + DIAB + CVD + HYPT + CANCER) #for unscaled regression reports#


#Model 2b (for AS315 only):
Same as above but only for DNAm collected from baseline screening visit


#Model 3:
lm(y ~ RaceEth + SEindex + Smoking + BMI + TEXPWK + HYST + DIAB + CVD + HYPT + CANCER + ALCOHOL + HEI2010_TOTAL_SC) #for unscaled regression reports#


#Explanation of variables:
#y are the age residualized DNAm readouts fitted separately
#RaceEth is self-reported race/ethnicty
#SEindex is socioeconomic index
#Smoking is 0 = never smoked, 1 = past smoker, 2 = current smoker
#BMI is body mass index
#TEXPWK is recreational energy expenditure per week
#HYST is history of hysterectomy; DIAB is history of diabetes; CVD is history of cardiovascular disease; HYPT is history of hypertension; CANCER is history of cancer
#ALCOHOL is Alcohol Intake 1: Non drinker, 2 is Past drinker, 3 is <1 drink per month, 4 is <1 drink per week, 5 is 1 to <7 drinks per week, 6 is 7+ drinks per week
#HEI2010 is the 2010 Total Healthy Eating Index Score


###########################################################
#########PART 4: Multivariable Cox Regression##############
###########################################################

##race/ethnicity stratified##

coxph(Surv(ENDFOLLOWALLDY, DEATHALL) ~ scale(Seindex) + scale(y) + Age + Smoking + BMI + DIAB + CVD + CANCER + HRTARM + DMARM + ALCOHOL + HEI2010_TOTAL_SCORE + strata(RaceEth), data=dat)


##separate analysis in the three race/ethnicity groups##

coxph(Surv(ENDFOLLOWALLDY, DEATHALL) ~ scale(Seindex) + scale(y) + Age + Smoking + BMI + DIAB + CVD + CANCER + HRTARM + DMARM + ALCOHOL + HEI2010_TOTAL_SCORE, data=subset(dat, RaceEth =="eth") #where "eth" is either White, Black, or Hispanic#


#Explanation of variables:
#ENDFOLLOWALLDY is Days enrollment to end of followup (including AllDiscovered Death)
#DEATHALL is All Discovered Death
#Age is chronological Age
#Smoking is 0 = never smoked, 1 = past smoker, 2 = current smoker
#BMI is body mass index
#DIAB is history of diabetes; CVD is history of cardiovascular disease; CANCER is history of cancer
#y is DNAm readout
#SEindex is socioeconomic index
#HRTARM is Hormone Replacement Therapy Arm
#DMARM is Dietary Modification Arm
#ALCOHOL is Alcohol Intake 1: Non drinker, 2 is Past drinker, 3 is <1 drink per month, 4 is <1 drink per week, 5 is 1 to <7 drinks per week, 6 is 7+ drinks per week
#HEI2010 is the 2010 Total Healthy Eating Index Score
#RaceEth is self-reported race/ethnicty
